# Clinical and pathological implications of the presence of MECA-79-expressing tumor cells in pathological stage IA lung adenocarcinoma

**DOI:** 10.1101/2025.04.08.25325444

**Authors:** Tomohito Saito, Mitsuaki Ishida, Tomoya O. Akama, Shiho Hattori, Natsumi Maru, Takahiro Utsumi, Aki K. Kobayashi, Kento J. Fukumoto, Hiroshi Matsui, Yohei Taniguchi, Yoshinobu Hirose, Katsuyasu Kouda, Tomohiro Murakawa

## Abstract

Approximately 15 % of patients with resected pathogenic stage IA lung adenocarcinoma develop recurrent disease, indicating the formation of a cancer metastasis-promoting microenvironment. MECA-79 epitope is a glycan structure modulating immune response, normally expressed on high endothelial venules. Recently, ectopic MECA-79 expression has been discovered in several cancer cells and was associated with poor prognosis. In this retrospective cohort study, we aimed to investigate the clinical and pathological significance of tumoral MECA-79 expression in early-stage lung cancer.

Using MECA-79 antibody to identify MECA-79^+^ tumor cells, we analyzed 195 patients with pathological stage IA lung adenocarcinoma undergoing lobectomy, assessing clinical, radiological, and pathological factors. We applied the Kaplan-Meier analysis to evaluate overall and recurrence-free survival as well as univariate and multivariate logistic regression to identify risk factors of postoperative recurrence within 5 years. Statistical significance was set at *P* < 0.05.

Among 195 patients undergoing lobectomy, immunohistochemical analysis revealed tumoral MECA-79 expression in 5.1 % of cases (n = 10). Patients with MECA-79^+^ tumor cells exhibited a larger pathological invasive size (2.1 vs. 1.6 cm, *P* = 0.044), along with higher vascular invasion rates (90.0 % vs. 40.0 %, *P* = 0.0023) and 5-year postoperative recurrence (40.0 % vs. 7.6 %, *P* = 0.0061). Kaplan-Meier analysis demonstrated significantly worse recurrence-free survival for patients with MECA-79^+^ tumor cells (5-year rate: 54.9 % vs. 87.4 %, *P* = 0.003). Multivariate logistic regression identified presence of MECA-79^+^ tumor cells as an independent predictor of 5-year postoperative recurrence (odds ratio, 6.51; *P* = 0.025).

Our results indicated that tumoral MECA-79 expression is associated with recurrence of resected pathological stage IA lung adenocarcinoma, warranting validation in multicenter cohorts.

## Introduction

Lung cancer is the leading cause of cancer-related deaths worldwide, with an estimated 1.8 million lung cancer-related deaths in 2022 [1]. Anatomic surgical resection is the mainstay definitive treatment for early-stage non-small cell lung cancer (NSCLC), and pathological stage IA disease accounts for approximately 50% of resectable NSCLC [2]. However, postoperative recurrence rate of pathological stage IA NSCLC remains as high as approximately 15% [3], which is equivalent to that of pathological stage II colorectal cancer [4]. Delineation of the underlying mechanism of such a high recurrence rate would lead to better clinical outcomes.

Sialyl Lewis^X^ (sLe^X^), an established serum tumor marker, is an important glycan epitope in tumor progression through selectin interactions. A sulfated derivative of sLe^X^, 6-sulfo sLe^X^, is also an important L-, P-, and E selectin ligand. Immunohistochemically, 6-sulfo sLe^X^ is recognized by MECA-79 antibody exclusively when 6-sulfo sLe^X^ is present on a particular glycan core structure, the MECA-79 epitope: 6-sulfo *N*-acetyllactosamine on the extended core 1 *O*-glycans, Galβ1→4(sulfo→6)GlcNAcβ1→3Galβ1→GalNAc→*O*-R [5]. In contrast to sLe^X^, 6-sulfo sLe^X^ as well as MECA-79 epitope have mainly been studied on non-malignant cells such as high endothelial venules in an immune modulation context. Recently, ectopic MECA-79 epitope expression in human cancer cells has been discovered and proven to be associated with poor prognosis in cholangiocarcinoma as well as gastric, bladder, and breast cancers [6–11]. However, to date, the presence of MECA-79^+^ tumor cells in NSCLC and the clinical and pathological significance of such presence have not been described.

The present study aimed to investigate clinical and pathological significance of the presence of MECA-79^+^ tumor cells in pathological stage IA lung adenocarcinoma (LUAD).

## Materials and methods

This study was conducted in accordance with the principles outlined in the Declaration of Helsinki as revised in 2013. This study was approved by the Kansai Medical University Hospital Research Ethics Committee (approval number: 2016662; approval date: September 12, 2016). The requirement for informed consent was waived as this retrospective study analyzed anonymized data and posed minimal risk to participants. Chart reviews were conducted on 2021 August 26th and 2024 December 31st for research purposes. At no point after the data collection did the authors have access to information that could identify individual participants. Archived pathological samples were collected and anonymized prior to analysis, and immunostaining and evaluation of these samples were performed between 2021 August 26th and 2021 October 29th.

### Patient selection

A flow diagram illustrating the patient selection process is shown in Fig 1. The records of 1014 patients who underwent surgery for lung cancer at Kansai Medical University Hospital between January 1, 2009, and December 31, 2017, were reviewed. The inclusion criteria were a pathological diagnosis of primary invasive LUAD, pathological stage IA (as per the eighth edition of the TNM classification system) [12], complete resection (R0) with lobectomy or greater, and tissue available for this study. The exclusion criteria were adenocarcinoma *in situ*, minimally invasive adenocarcinoma, surgery for biopsy/diagnosis, and patients who had received neoadjuvant therapy. Based on these criteria, 195 patients who had undergone pulmonary lobectomy with lobe-specific lymph node dissection were included for analysis in this study.

**Fig 1.**
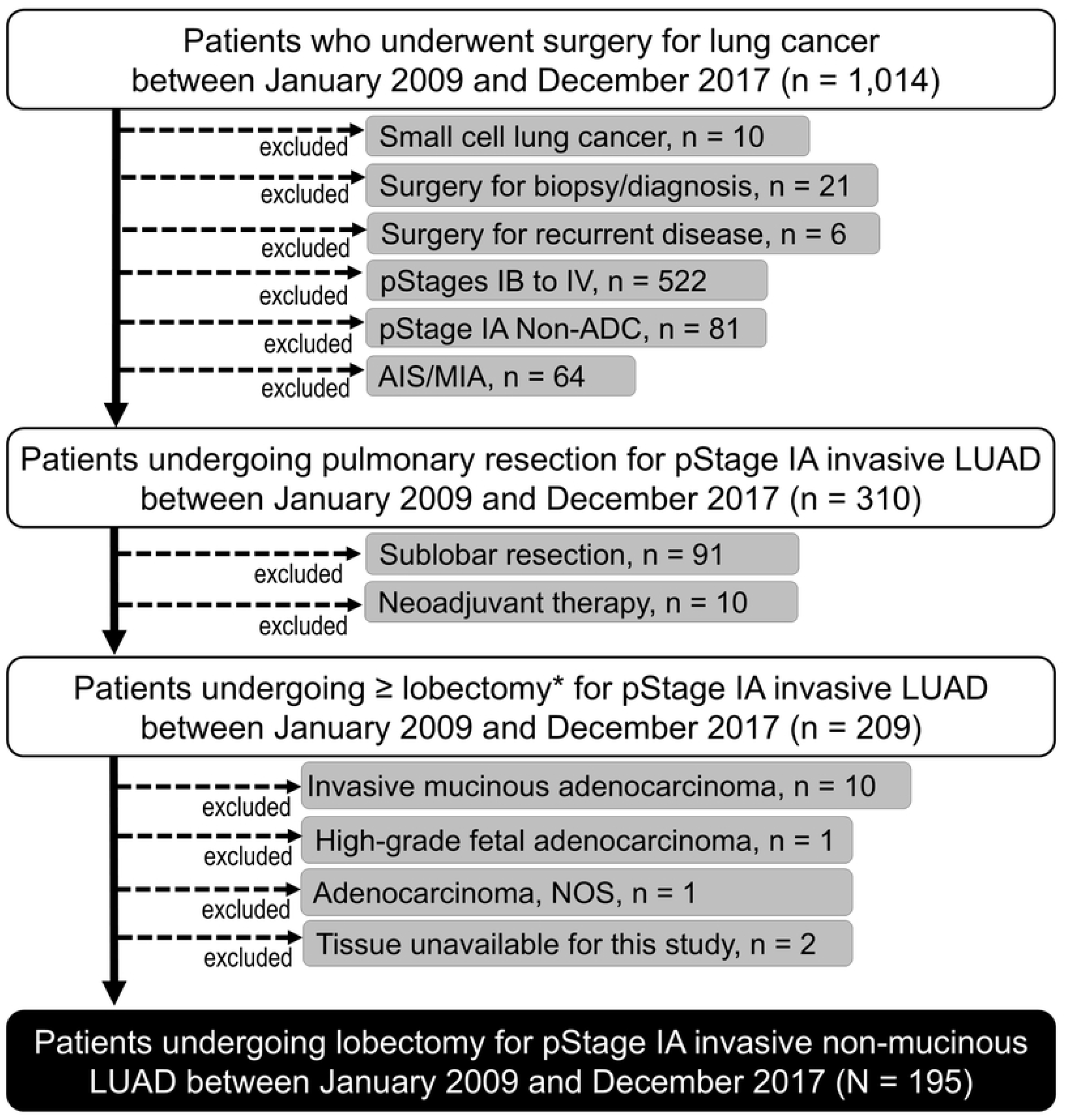
Flow Diagram of the Patient Selection Process. Briefly, out of 564 patients undergoing surgery for lung cancer at Kansai Medical University Hospital between January 1, 2009 and January 31, 2015, a total of 100 patients who underwent complete resection with lobectomy for pathological stage IA invasive adenocarcinoma of the lung with no neoadjuvant therapy were included in this study. Adenocarcinoma *in situ* and minimally invasive adenocarcinoma were excluded.

### Diagnosis of invasive LUAD

The resected specimens were handled in accordance with the Japan Lung Cancer Society General Rule for Clinical and Pathological Records of Lung Cancer [13]. The pathological diagnosis of invasive LUAD was made in accordance with the 2015 World Health Organization (WHO) histological classification of lung tumors [14]. Grading was determined according to The International Association for Study of Lung Cancer (IASLC) grading system for invasive LUAD based on a recent report [15]. In addition to vascular invasion and lymphatic permeation, spread through air spaces (STAS) was evaluated as previously described [16].

### Immunohistochemical analyses

Formalin-fixed, paraffin-embedded tissues cut into 5-μm-thick sections were used for hematoxylin-eosin staining and for immunohistochemical analysis using MECA-79 antibody (1:100; MECA-79; Novus Biologicals, Englewood, CO, USA). Tumoral expression of MECA-79 was reviewed by a trained pathologist (MI). Representative microscopic images of MECA-79^+^ tumor cells in this study are shown in Fig 2.

**Fig 2.**
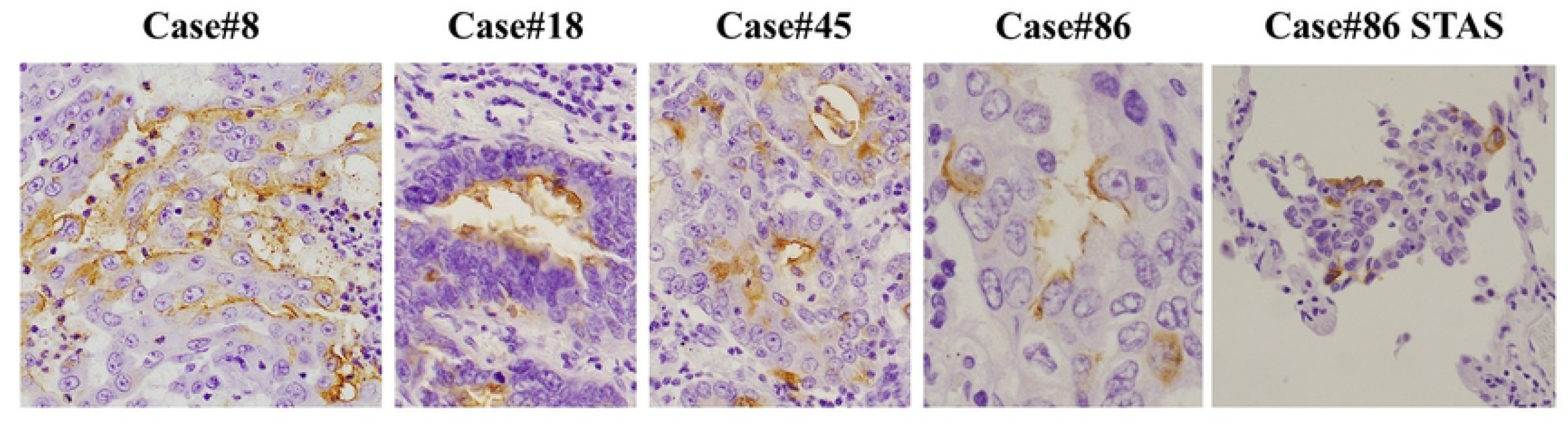
Representative Microscopic Images of Tumoral MECA-79 Expression in Human Lung Adenocarcinoma Cells Tumoral expression of MECA-79 was observed predominantly in the cell membrane and occasionally in the cytoplasm. In case #86, MECA-79^+^ tumor cells were also found in cell clusters of spread through air spaces (STAS). MECA-79 antibody (Novus Biologicals, Englewood, CO, USA) was used for immunostaining.

### Radiological features on chest computed tomography (CT) and 18-fluorodeoxyglucose positron emission tomography

Preoperative chest computed tomography (CT) images were obtained with a slice thickness of 1–10 mm and a field of view of 14–39 cm. The type of pulmonary nodule (solid, part-solid, or pure ground-glass nodule) and the size of the nodule (total size and solid size) were determined only in patients who underwent high-resolution CT with a slice thickness of 1–2 mm, in accordance with previous reports [17,18]. For patients who had undergone preoperative 18-fluorodeoxyglucose positron emission tomography/CT and whose radiological diameter of the pulmonary nodule was ≥ 1.0 cm, the maximum standardized uptake value within the primary lesion was recorded.

### Statistical analysis

Differences in non-parametric continuous and categorical variables were determined using the Kruskall-Wallis and chi-square tests, respectively. Postoperative recurrence of LUAD was diagnosed considering the evidence provided by physical examination and diagnostic imaging such as CT, MRI, and FDG-PET/CT as previously described [19]. Recurrent disease was pathologically confirmed when clinically feasible. The Kaplan-Meier estimation curve was used to visually analyze overall survival (OS) and recurrence-free survival (RFS), and the difference between groups was evaluated using the log-rank test. OS was calculated as the interval between the date of pulmonary resection and the date of all-cause death. RFS was calculated as the interval between the date of pulmonary resection and the date of first documented recurrence or all-cause death. The date of the last follow-up was used for censored patients.

Univariate and multivariate logistic regression analyses were used to evaluate the association of clinical and pathological characteristics with postoperative recurrence of LUAD within 5 postoperative years. Factors that were statistically significant (*P* < 0.05) in the univariate analysis were considered as candidate risk factors. First, all candidate factors were included in the multivariate analysis. Second, given the 17 postoperative recurrences observed within 5 postoperative years (as described below), two independent variables, including the presence of MECA-79+ tumor cells, were selected for multivariable model construction. This approach adhered to the general rule requiring at least 10 outcomes per independent variable in logistic regression analysis.[20]

Data was analyzed using JMP Pro version 18.2.0 (JMP Statistical Discovery LLC, NC, USA) for Windows (Microsoft, Redmond, WA, USA). The Kaplan-Meier estimation curve was visualized using Prism version 6.0.7 (GraphPad Software, MA, USA). Statistical significance was set at *P* < 0.05.

## Results

The clinical and pathological characteristics of the study population according to the presence or absence of MECA-79^+^ tumor cell are summarized in Table 1. Of the 195 patients included for analysis, ten (5.1%) showed MECA-79^+^ tumor cells. Radiological solid and pathological invasive sizes were slightly greater in patients with MECA-79^+^ tumor cells than in their counterparts (*P* = 0.046 and 0.044, respectively). Vascular invasion and recurrence within 5 postoperative years were more commonly observed in patients with MECA-79^+^ tumor cells (*P* = 0.0023 and 0.0061, respectively). The presentation of postoperative recurrence was not significantly different between the two study groups (*P* = 0.77).

**Table 1.**
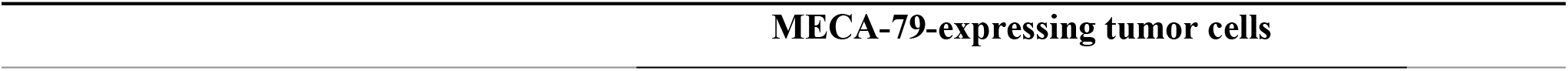
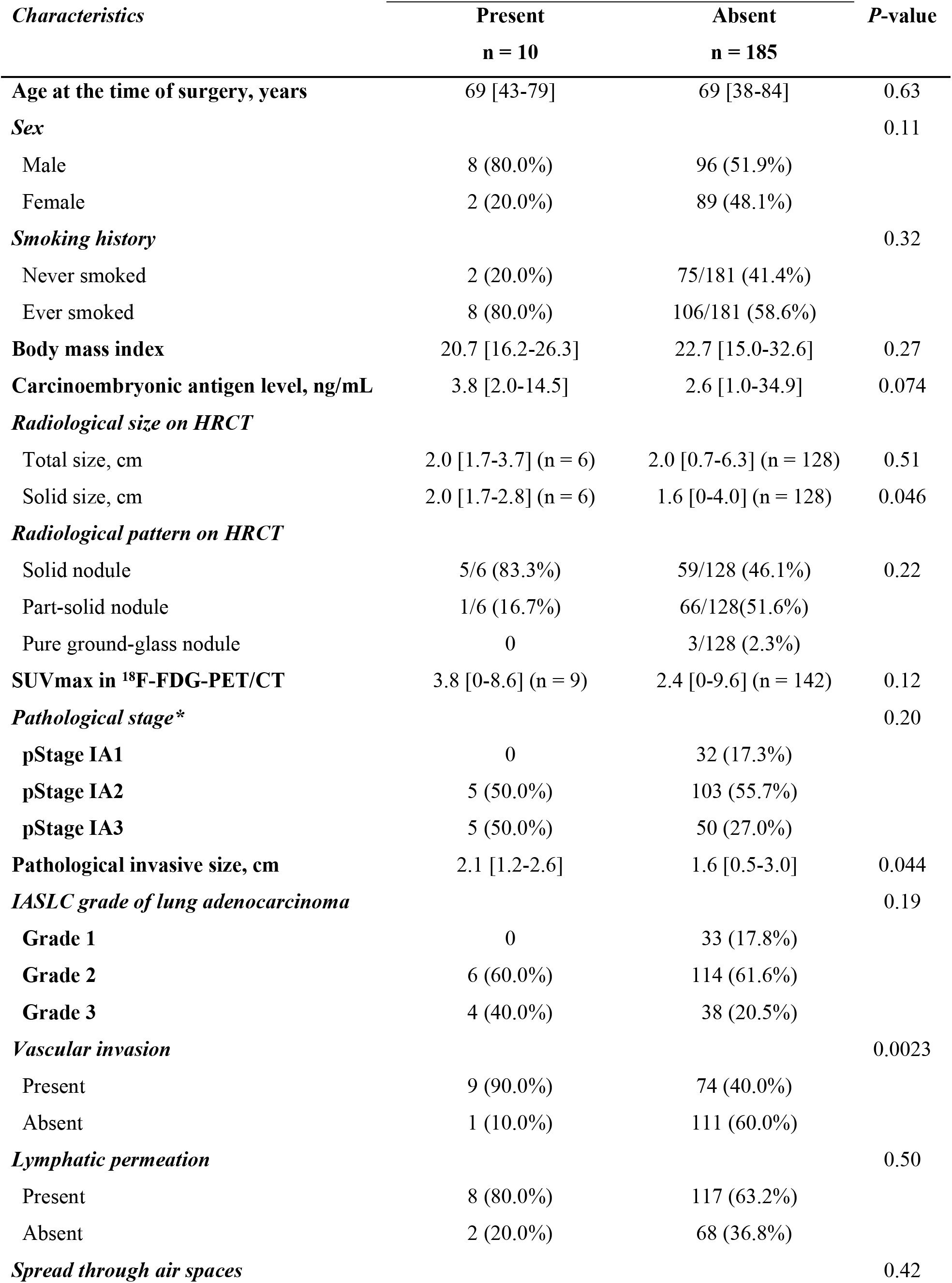
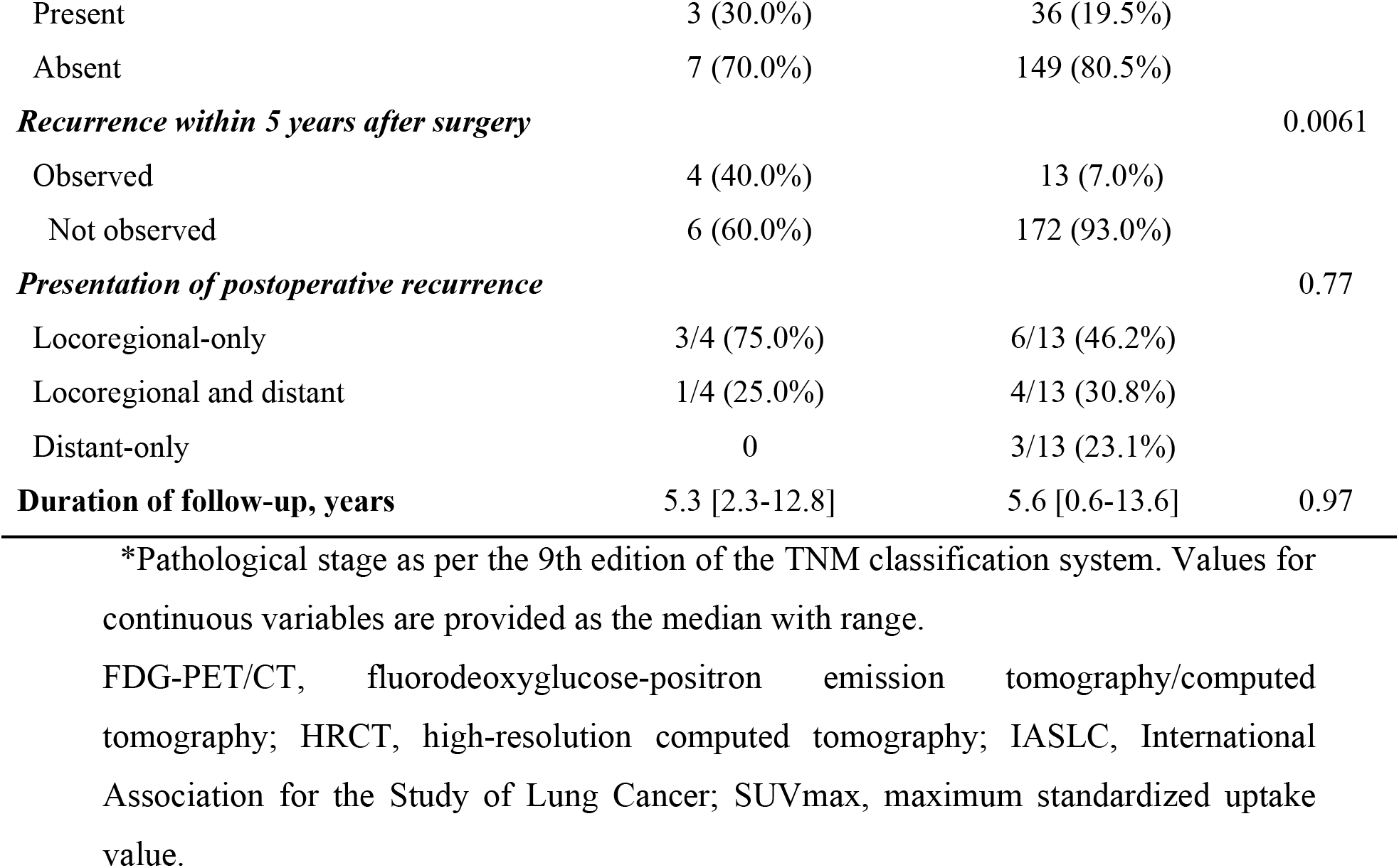
Baseline Clinical and Pathological Characteristics of Patients Undergoing Lobectomy for Pathological Stage IA Lung Adenocarcinoma according to Tumoral MECA-79 Status (N = 195).

| *Characteristics* | **Present**<br>**n = 10** | **Absent**<br>**n = 185** | *P-value* |
| --- | --- | --- | --- |
| **Age at the time of surgery, years** | 69 [43-79] | 69 [38-84] | 0.63 |
| **Sex** |  |  | 0.11 |
| Male | 8 (80.0%) | 96 (51.9%) |  |
| Female | 2 (20.0%) | 89 (48.1%) |  |
| **Smoking history** |  |  | 0.32 |
| Never smoked | 2 (20.0%) | 75/181 (41.4%) |  |
| Ever smoked | 8 (80.0%) | 106/181 (58.6%) |  |
| **Body mass index** | 20.7 [16.2-26.3] | 22.7 [15.0-32.6] | 0.27 |
| **Carcinoembryonic antigen level, ng/mL** | 3.8 [2.0-14.5] | 2.6 [1.0-34.9] | 0.074 |
| **Radiological size on HRCT** |  |  |  |
| Total size, cm | 2.0 [1.7-3.7] (n = 6) | 2.0 [0.7-6.3] (n = 128) | 0.51 |
| Solid size, cm | 2.0 [1.7-2.8] (n = 6) | 1.6 [0-4.0] (n = 128) | 0.046 |
| **Radiological pattern on HRCT** |  |  |  |
| Solid nodule | 5/6 (83.3%) | 59/128 (46.1%) | 0.22 |
| Part-solid nodule | 1/6 (16.7%) | 66/128 (51.6%) |  |
| Pure ground-glass nodule | 0 | 3/128 (2.3%) |  |
| **SUVmax in <sup>18</sup>F-FDG-PET/CT** | 3.8 [0-8.6] (n = 9) | 2.4 [0-9.6] (n = 142) | 0.12 |
| **Pathological stage\*** |  |  | 0.20 |
| pStage IA1 | 0 | 32 (17.3%) |  |
| pStage IA2 | 5 (50.0%) | 103 (55.7%) |  |
| pStage IA3 | 5 (50.0%) | 50 (27.0%) |  |
| **Pathological invasive size, cm** | 2.1 [1.2-2.6] | 1.6 [0.5-3.0] | 0.044 |
| **IASLC grade of lung adenocarcinoma** |  |  | 0.19 |
| Grade 1 | 0 | 33 (17.8%) |  |
| Grade 2 | 6 (60.0%) | 114 (61.6%) |  |
| Grade 3 | 4 (40.0%) | 38 (20.5%) |  |
| **Vascular invasion** |  |  | 0.0023 |
| Present | 9 (90.0%) | 74 (40.0%) |  |
| Absent | 1 (10.0%) | 111 (60.0%) |  |
| **Lymphatic permeation** |  |  | 0.50 |
| Present | 8 (80.0%) | 117 (63.2%) |  |
| Absent | 2 (20.0%) | 68 (36.8%) |  |
| **Spread through air spaces** |  |  | 0.42 |
| Present | 3 (30.0%) | 36 (19.5%) |  |
| Absent | 7 (70.0%) | 149 (80.5%) |  |
| ***Recurrence within 5 years after surgery*** |  |  | 0.0061 |
| Observed | 4 (40.0%) | 13 (7.0%) |  |
| Not observed | 6 (60.0%) | 172 (93.0%) |  |
| ***Presentation of postoperative recurrence*** |  |  | 0.77 |
| Locoregional-only | 3/4 (75.0%) | 6/13 (46.2%) |  |
| Locoregional and distant | 1/4 (25.0%) | 4/13 (30.8%) |  |
| Distant-only | 0 | 3/13 (23.1%) |  |
| **Duration of follow-up, years** | 5.3 [2.3-12.8] | 5.6 [0.6-13.6] | 0.97 |

\*Pathological stage as per the 9th edition of the TNM classification system. Values for continuous variables are provided as the median with range. FDG-PET/CT, fluorodeoxyglucose-positron emission tomography/computed tomography; HRCT, high-resolution computed tomography; IASLC, International Association for the Study of Lung Cancer; SUVmax, maximum standardized uptake value.

While OS showed no significant difference between the two study groups (*P* = 0.32, Fig 3A), RFS was significantly worse for patients with MECA-79^+^ tumor cells than those with no MECA-79^+^ tumor cells (*P* = 0.003, Fig 3B).

**Fig 3.**
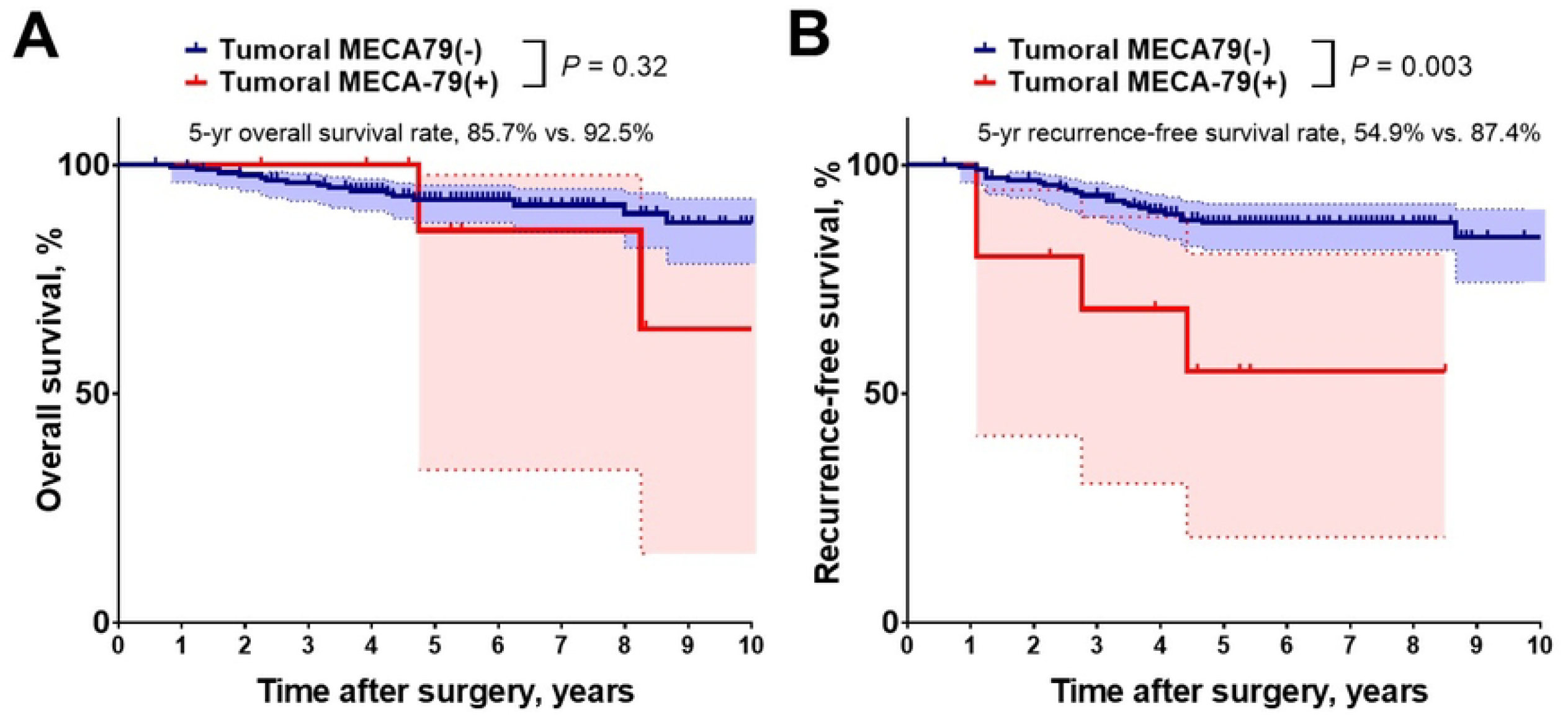
Overall and Recurrence-free Survival According to Tumoral MECA-79 Expression Status (A) Overall survival was not significantly different regardless of tumoral MECA-79 expression status. (B) Recurrence-free survival was significantly worse for patients with MECA-79^+^ tumor cells than for those with no MECA-79^+^ tumor cells (5-year recurrence-free survival, 54.9% vs. 87.4%, respectively, *P* < 0.001).

Univariate logistic regression analysis identified sex, smoking status, IASLC grade of LUAD, vascular invasion, lymphatic permeation, STAS, and presence of MECA-79^+^ tumor cells as candidate factors to predict postoperative recurrence within 5 years of surgery (Table 2). Multivariate logistic regression model including all candidate factors showed the presence of MECA-79^+^ tumor cells as an independent risk factor of postoperative recurrence within 5 years of surgery (Table 2). Further, the presence of MECA-79^+^ tumor cells was indicated as an independent risk factor of recurrence within 5 postoperative years in each multivariate logistic regression model including two selected candidate factors (Table 3).

**Table 2.**
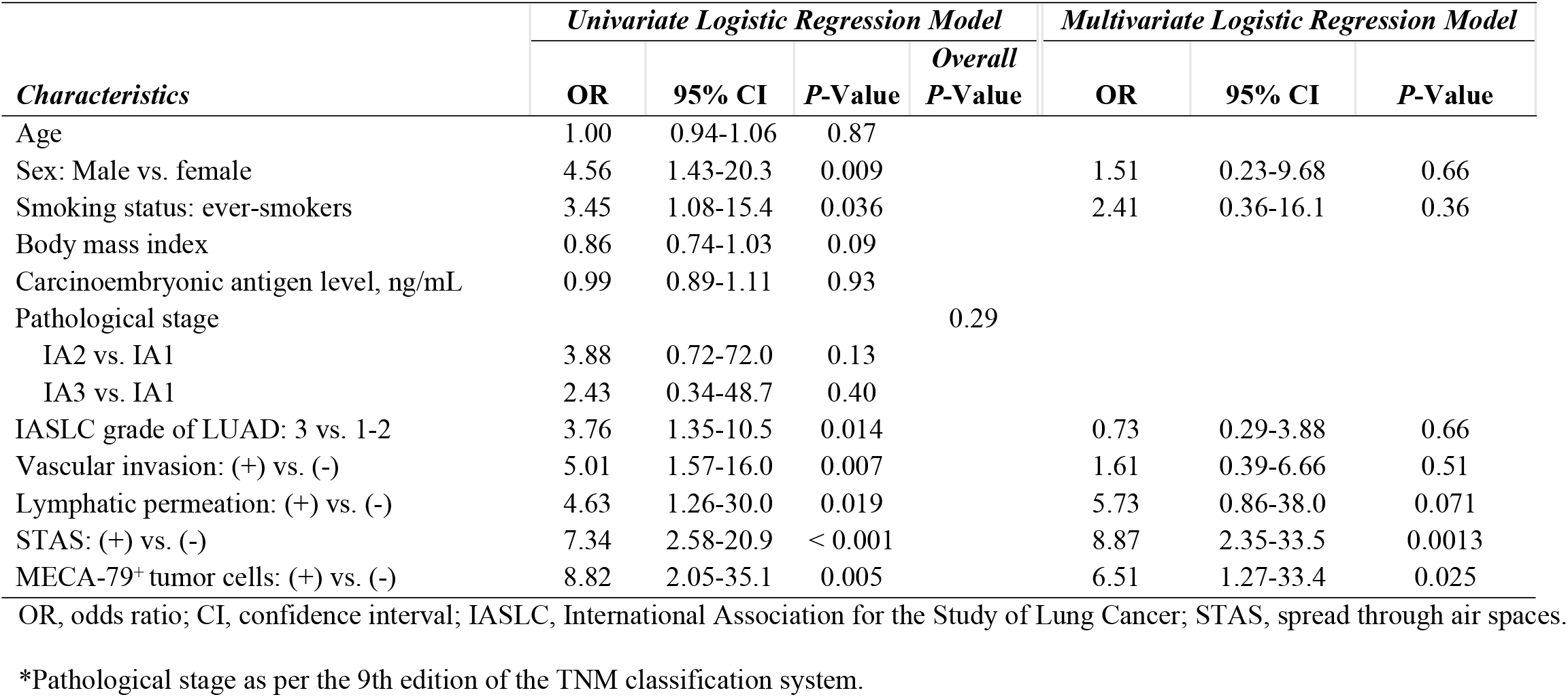
Univariate and Multivariate Logistic Regression Analysis on Clinical and Pathological Characteristics for Predicting Postoperative Recurrence within 5 Years of Surgery.

**Table 3.** Multivariate Logistic Regression Models Including Two Selected Candidate Factors for Postoperative Recurrence within 5 Years of Surgery.

| *Characteristics* | *Multivariate Model 1* |  |  | *Multivariate Model 2* |  |  | *Multivariate Model 3* |  |  |
| --- | --- | --- | --- | --- | --- | --- | --- | --- | --- |
|  | OR | 95% CI | P-Value | OR | 95% CI | P-Value | OR | 95% CI | P-Value |
| Sex: Male vs. female | 0.25 | 0.06-0.93 | 0.039 |  |  |  |  |  |  |
| Smoking status: ever-smokers |  |  |  | 3.06 | 0.83-11.3 | 0.093 |  |  |  |
| IASLC grade of lung adenoca.:<br>3 vs. 1-2 |  |  |  |  |  |  | 3.38 | 1.17-9.80 | 0.025 |
| Vascular invasion: (+) vs. (-) |  |  |  |  |  |  |  |  |  |
| Lymphatic permeation: (+) vs. (-) |  |  |  |  |  |  |  |  |  |
| STAS: (+) vs. (-) |  |  |  |  |  |  |  |  |  |
| MECA-79<sup>+</sup> tumor cells: (+) vs. (-) | 6.93 | 1.67-28.8 | 0.008 | 7.43 | 1.81-30.5 | 0.005 | 7.61 | 1.80-32.1 | 0.006 |
| *Characteristics* | *Multivariate Model 4* |  |  | *Multivariate Model 5* |  |  | *Multivariate Model 6* |  |  |
|  | OR | 95% CI | P-Value | OR | 95% CI | P-Value | OR | 95% CI | P-Value |
| Sex: Male vs. female |  |  |  |  |  |  |  |  |  |
| Smoking status: ever-smokers |  |  |  |  |  |  |  |  |  |
| IASLC grade of lung adenoca.:<br>3 vs. 1-2 |  |  |  |  |  |  |  |  |  |
| Vascular invasion: (+) vs. (-) | 3.94 | 1.18-13.1 | 0.025 |  |  |  |  |  |  |
| Lymphatic permeation: (+) vs. (-) |  |  |  | 4.26 | 0.93-19.6 | 0.063 |  |  |  |
| STAS: (+) vs. (-) |  |  |  |  |  |  | 7.69 | 2.55-23.1 | < 0.001 |
| MECA-79<sup>+</sup> tumor cells: (+) vs. (-) | 5.27 | 1.25-22.2 | 0.024 | 7.90 | 1.91-32.7 | 0.004 | 9.70 | 2.01-45.7 | 0.004 |

OR, odds ratio; CI, confidence interval; IASLC, International Association for the Study of Lung Cancer; STAS, spread through air spaces.

## Discussion

Our retrospective immunohistochemical analysis of 195 resected pathological stage IA LUAD cases indicated that the presence of MECA-79^+^ tumor cells might be associated with postoperative recurrence within 5 years after surgery and with vascular invasion, but neither with lymphatic permeation nor STAS.

To date, there have been six reports on tumoral expression of the MECA-79 epitope in human cancer: gastric cancer [6], cholangiocarcinoma [7–9], bladder cancer [10], and breast cancer [11]. The proportion of patients showing MECA-79^+^ tumor cells seem to vary by cancer types: 28% for gastric cancer [6], 42%–70% for cholangiocarcinoma [7,9], 20% for bladder cancer [10], and 0% for hepatocellular carcinoma [7]. In gastric cancer, the presence of MECA-79^+^ tumor cells seems to be associated with venous invasion, distant metastasis, and poor cancer-specific survival in gastric cancer. In our study, we observed a relatively low MECA-79^+^ tumor-cell positivity rate and associations of the presence of MECA-79^+^ tumor cells with vascular invasion and postoperative recurrence in LUAD, while we did not find a significant increase in distant metastasis in patients with MECA-79^+^ tumor cells.

The pathophysiological mechanism linking tumoral MECA-79 expression and cancer recurrence remains to be fully elucidated. Given that the MECA-79 epitope comprises a part of 6-sulfo sLe^X^, the pro-metastatic roles of tumoral 6-sulfo sLe^X^ expression could explain the increase in vascular invasion and cancer recurrence observed in the presented study. Tumoral 6-sulfo sLe^X^ has been shown to bind E-selectin on vascular endothelial cells via calcium-dependent interactions, facilitating vascular invasion in bladder cancer [10]. Further, tumoral 6-sulfo sLe^X^ expression enables cancer cells to form microthrombi with platelets via 6-sulfo sLe^X^ binding to P-selectin, which may create protective niches that evade immunological surveillance [21]. However, the anti-metastatic role of tumoral sLe^X^ has also been shown in oral squamous cell carcinoma [22]. Tumoral sLe^X^ can attract CD8+ cytotoxic T lymphocytes, promoting the formation of immune synapses. Another possibility could be that tumoral MECA-79 expression is phenotypic rather than functional and that the underlying mechanism of upregulation of MECA-79 in cancer cells promotes cancer recurrence. For example, overexpression of beta-1,3-N-acetylglucosaminyltransferase, an essential enzyme for the biosynthesis of MECA-79, has been reported to be associated with diminished disease-free and OS and with increased infiltration of regulatory T cells [23–25].

The limitations of the presented study were as follows: first, our results are based on a retrospective single-institutional study with a small sample size. Thus, patient characteristics might have influenced our data and we acknowledge limited generalizability. A prospective, multi-institutional study with a larger sample size is needed to validate the current results. Second, this study only included pathological stage IA LUAD. Thus, the positivity rate and clinical significance of the presence of MECA-79^+^ tumor cells in patients with pathological stages IB–IV, with neoadjuvant or adjuvant therapy, and with non-adenocarcinoma non-small cell lung cancer should be investigated in further studies. Third, we did not have background data on gene alterations such as epidermal growth factor receptor, Kirsten rat sarcoma viral oncogene homolog, and anaplastic lymphoma kinase mutations. Thus, the relationship between tumoral MECA-79 expression and the genetic background will need to be clarified in future studies.

In conclusion, our results indicate that the presence of MECA-79^+^ tumor cells could be associated with vascular invasion and postoperative recurrence of resected pathological stage IA LUAD. Further investigations are necessary to validate our results and delineate the underlying mechanisms.

## Data Availability

All relevant data are within the manuscript and its Supporting Information files.

## Acknowledgements

This work was supported by JSPS KAKENHI Grant # JP20K09184 and # JP24K12309. The authors would also like to thank Editage (www.editage.com) for English language editing.

